# Rapid increase of Care Homes reporting outbreaks a sign of eventual substantial disease burden

**DOI:** 10.1101/2020.05.07.20089243

**Authors:** Ian Hall, Hugo Lewkowicz, Thomas House, Lorenzo Pellis, James Sedgwick, Nick Gent

## Abstract

Enclosed societies (i.e. locations that are connected to wider community only by subgroups of their population and that are dominated by within society transmission) have the potential, upon establishment of a respiratory disease, to suffer a large proportion of the population within becoming infected. Care homes are particularly susceptible to COVID19 outbreaks and suffer high mortality due to vulnerable population within. Recent data on the number of new outbreak reports in care homes to Public Health England shows an initial increase then plateau perhaps associated with an SIS model dynamic. Without change in policy moving forward a high prevalence in such setting is predicted of around 75%. Action is needed to support staff in such settings.

---

Enclosed societies have the potential, upon establishment of a respiratory disease, to suffer a large proportion of the population within becoming infected. Early reports of a novel infection circulated in December 2019 and in January 2020 attributed to a novel coronavirus, SARS-CoV2. Since then, the disease has become a pandemic with documented outbreaks in most countries.^1^ Outbreaks have been reported in many settings. The outbreak of COVID-19 on board the cruise ship Diamond Princess first came to attention when Japanese authorities quarantined the vessel in Yokohama on the 5th February 2020.^2^ Prisons and care homes share similarities to cruise ships conceptually in terms of having a stable resident population and a staff population. However, in those settings the staff population is much better connected to a wider community. Internationally there have been a number of outbreaks in care homes with substantial associated attack rates and mortality.^3^

Reviews of outbreaks of influenza in enclosed societies have shown that while the particular kind of society (prison, care home, school, barracks, etc.) was not a significant explanatory variable of attack ratio, a person’s occupation within the society was.^4,5^

The number of new reported outbreaks of COVID19 in care homes rose rapidly in early to mid-March. Since late-March the number reported each has plateaued, see Figure 1. There is a clear weekend lull in reporting outbreaks. The early rapid growth in new reported outbreaks is consistent with the exponential growth expected from epidemic theory. Indeed it is this rapid growth in unmitigated outbreaks that is a major cause of concern.^6^

**Figure 1:**
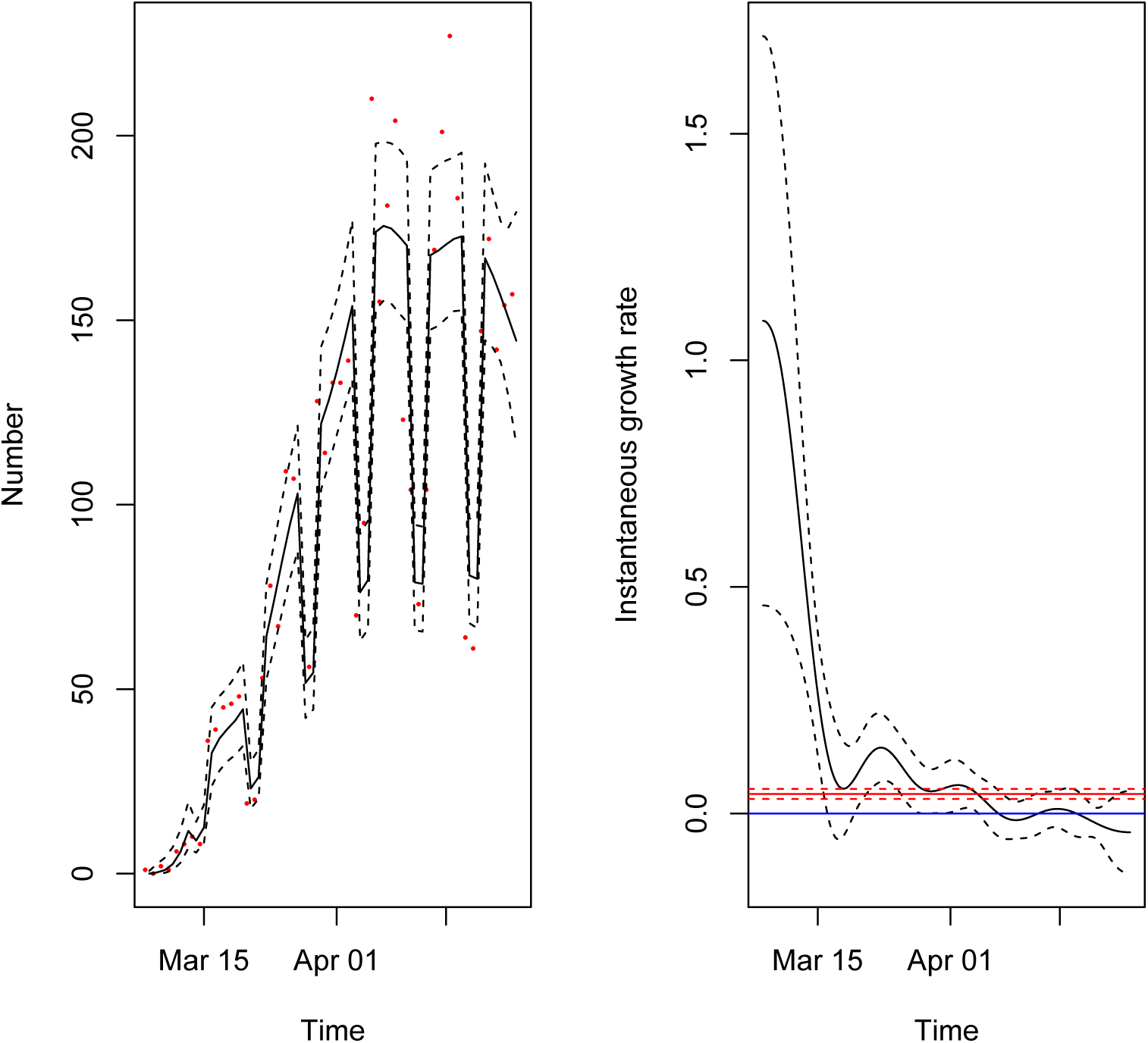
Left panel: Number of care home outbreaks reported each day to Public Health England^7^ (red dots), Generalised Additive Model (GAM) best fit curve (black line) and CI (dashed lines) (Quasi-Poisson link with spline on time and fixed effect on weekend or weekday); Right panel: Instantaneous growth rate in number of reported care homes in England from GAM (black) and Generalised Linear Model (red, Quasi-Poisson link without day of week explanatory variable)

We fit a generalised additive model to the observed new reported outbreaks with a fixed effect variable for weekend and weekday and a spline fitted to time. The observations are plotted as red points on left hand panel, the black line shows the corresponding best fit and dashed black line the 95% confidence interval on the estimate.

The right panel shows the derivative of the spline arising from the GAM. If this derivative is constant and positive then this is a sign of exponential growth, if constant and negative of exponential decay, and if it is tracking the blue line (a reference line for zero growth) this is suggestive that the data is plateauing. The model seems to be suggesting 3 phases of growth in the data, initially very rapid growth followed by a couple of weeks of slower growth followed by 2 weeks of plateauing. The red line on this graph shows the growth rate estimated from a simple generalised linear model to whole data set for comparison.

This data shows the number of care homes reporting an outbreak of COVID19, but not the state of the outbreak within each care home. As a result, traditional epidemic modelling is not possible. Using an ’SIS’ model with constant force of infection into each care home (as an infectious unit rather than individuals) to model the outbreak across care homes does not explain the data well. However this is probably owing to a time varying transmission term. The spline used in the GAM above is a proxy for this mathematical form and given that SIS theory predicts a steady state of constant fraction of care homes in outbreak status we project the current trend observed into the future

We may infer the eventual prevalence of outbreak status care homes if this trend continues which would be *P_∞_* = *TQ_∞_* where *T* is the recovery time, *Q_∞_*, is the fraction of the care homes in England that report each day at stead state incidence. According to CQC, the number of care homes is *N* = 15517.^8^ We find that an upper limit estimate on *Q_∞_*, = 190/*N* from the two week plateau seen in the data and fit by the statistical model which we use as a worst case scenario. The recovery time of a care home is uncertain. If the generation time is taken to be 5 days^9^ and 3-4 generations of disease occur with associated wait of a couple of 14 days after last observed case then *T* ~ 34 is plausible meaning that *P_∞_* = 0.41. If *T* = 60 days (allowing for weaker infection control and undiagnosed cases) then *P_∞_* = 0.73 is potential reasonable worst case planning end state.

This analysis is relatively simple and does not inform the size of the outbreaks within homes themselves. This will be variable based on local outbreak management. However, it is important to observe that the 73% value is the prevalence of care homes with outbreaks and not the incidence. This means homes should expect to suffer multiple importations over time. These outbreaks will accumulate cases so the final attack rate within a home may be large due to a mix of explosive outbreaks and repeated importation.

The early growth in the data that reduces the ability of constant force of infection explanations may be an artefact of surveillance (a relatively new scheme having improved uptake in usage over time) rather than epidemiology but the plateauing numbers of reports is still relatively substantial. Additionally the route of infection importation may be higher in early stages from the community (visitors) or from cases being discharged from hospital. Indeed early on we should expect the generation of new cases to track the community outbreak. These processes should have been stopped once social distancing interventions were in place but may explain the early fast growth and the later plateau may be a sign of a change in transmission term varying over time. In the UK current intervention were implemented on 24th March 2020, after which the spline appears to be more stable.

A plausible explanation of the model presented in this correspondence is that the vehicle of connecting care homes is the staff. Staff seem to be suffering disease at similar numbers to residents (it should be noted that the reason for staff absence is unclear in the data and may be that staff are absent for precautionary reasons or care of dependents elsewhere, rather than due to the disease directly). If staff work in multiple care homes then these high attack rates may lead to depletion of susceptible staff and so reduce transmission in time. However, recent statements from WHO suggests evidence for long lasting immunity is limited.^10^ Moreover, staff interact with households and community and other care homes, so infection can be passed between care homes with staff acting as main vectors. This is not a sign that staff are being unobservant of the current severity of the situation but highlights the challenges they face and the high numbers of mild symptoms in cases.

The fact that the number of new reports has dipped in past week may be a sign of changing practises in some areas (the apparent reduction is driven by a single region of England that suffered a large number of outbreaks). This suggests that further investigation is imperative in understanding the outbreaks within care homes, explicitly including the role of staff. This can be established through enhanced testing and monitoring the interactions of care homes with the with wider community. It also suggests the need for support intervention to assist outbreak management and hard pressed staff working in the social care sector.

## Data Availability

Data is being published online on weekly basis.

https://www.gov.uk/government/statistical-data-sets/covid-19-number-of-outbreaks-in-care-homes-management-information

## Supplementary Material

### Data

The data, reported to Public Health England by its network of Health Protection Teams, show the number of new care homes reporting an outbreak of at least one case each day, starting from 8th March 2020 to 24th April 2020. A weekly variant is published online by PHE.^7^

### Statistical analysis

Using a Generalised Additive Model to look at trend over time (in R using mgcv with quasi-Poisson link)^11^ in England, Figure 1, shows signs of plateau in reported outbreaks since early April. For the first 20 days or so there was rapid growth with a 3-4 day doubling time.

Dips in reporting at weekends can be seen and a week day/ weekend term was included as a fixed effect explanatory variable alongside day of report which is fitted with a spline. This shows evidence of plateauing over past couple of weeks with a growth rate likely to be zero.

Whilst the most recent data points used are slightly lower, this is possibly due to regional variation and the reduction is not statistically significant.

### Mathematical Modelling

We assume care homes are in two states, either showing no cases of infectious disease or with some cases detected. Given the number of care homes *N* is fixed we can consider just the number of care homes reporting at lease one case, rather than the proportion.

The number of care homes with at least one case, *I*, will increase by further cases being detecting in previously unaffected care homes. It will decrease when laboratory confirmation arrives that the infected cases are not COVID-19 (fast timescale of a couple of days) or when the care home outbreak is declared over. Once declared infection free a care home may be reinfected later in pandemic.

Assuming an average rate *γ* can be constructed as a composite of the fast acting timescale of laboratory confirmation and slower timescale of outbreak cessation then *I → I −* 1 at some rate *γI*.

The increase in *I* is harder to model accurately. We assume there is a between care home infection rate *β* so that *I* → *I* + 1 with rate Λ(*N* — *I*) where *N* is the number of care homes in total.

A key question is what is an appropriate form of Λ

### Force of infection constant: Λ = λ

If the force of infection is constant then we may derive an ordinary differential equation (ODE) of the form

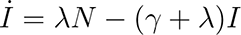

which has solution

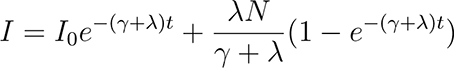

which will tend to a constant, predicting that a fraction λ/(*γ* + λ) of care homes will become affected.

However, the new reported affected care homes

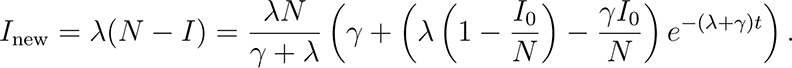

Whilst the data is currently plateauing according to the previous statistical analysis, the reported data in early stages of time series does not grow linearly as would be predicted by this model (rather the model predicts linear decay) so further refinement is necessary.

### Force of Infection proportional to infected care homes: Λ = β I/N

The force of infection Λ is likely not simply a constant. We can assume, instead, that the force of infection is proportional to the number of care homes currently suffering an outbreak (Λ = *β I*/*N*). This is an approximation. Clearly a physical home cannot infect another, but staff working between care homes, living and interacting socially with other staff, or doing multiple essential jobs may pass infection between. Essential transfers to/from hospitals or infected visitors (prior to lockdown) may have contributed.

The model constructed above can be written in a deterministic *SIS* ODE. This means we can write down an explicit solution for *I*:

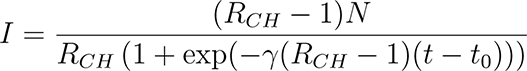

where *R_CH_* = *β*/*γ* and *t*_0_ = (ln (*R_CH_* (*N − I_0_*) − *N*) − ln (*R_CH_I_0_*))/(*γ*(*R_CH_* − 1)) controls behaviour of equation initially (assuming the number of infected care homes at time zero is *I*_0_) such that when *t* = *t*_0_ we have *I* half the value it will attain eventually. We note that *I* can be written in the form

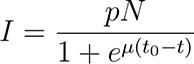

where 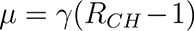, 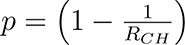 (the fraction of care homes infected at steady state). When *R_CH_* ≫ 1 this would suggest simple logistic regression is suitable or when *t* ≪ *t*_0_ we can assume exponential growth (log-linear regression).

However the data presented shows new care homes reported each day with no information about cessation of outbreaks. Thus in SIS framework this data would be those entering the *I* state namely

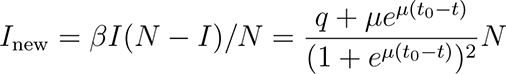

where *μ* and *t*_0_ are defined as above and *q* = *pγ* = *μ*_1_/*R_CH_* and we can extract the eventual prevalence *p* = *q*/*γ* = 1 − 1/*R_CH_*.

This model may be fitted to the data, and plausible fits are attained when *μ* ≪ *q*. However, when this is not the case the model structure loses its flexibility to fit as this second term is large.

Neither simple framework predicts new reported cases. We can attempt more complex models with time varying coefficients or variants on the form of Λ.

### Force of Infection time varying: Λ = γρ(t)I/N

In this case we are assuming a time varying trend on transmission term, but a fixed recovery rate

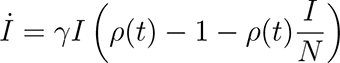

Making the transform *I*(*t*) = *P_∞_N*/(1 + *X*(*t*)) for some constant *P_∞_* we find

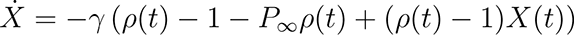

meaning that

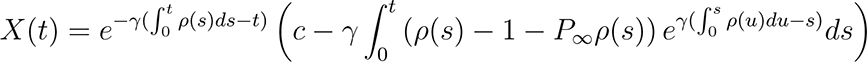

where *c* = *P_∞_N*/*I*_0_ − 1 defines the initial condition and that

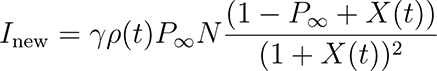

We could derive a form of *ρ*(*t*) to fit to the data, but we already have an estimate of *ρ*(*t*) in the form of the spline from the generalised additive model given suitable transformation.

We may choose the form of *ρ*(*t*) to evolve over time to be a constant (so that lim*_t→∞_ ρ*(*t*) = *R* for some *R* > 1 meaning that eventually *P_∞_* = lim*_t→∞_ I*(*t*) /*N* =1 − 1/*R*). In this case *R* may be viewed as the reproduction number in traditional epidemic modelling.

Projecting the model forward assumes of course the time varying transmission term *ρ*(*t*) is not affected by interventions in the future and plateau remains stable. Clearly if further outbreak control is effective the transmission term should reduce so that new outbreaks reduce in the future. In terms of planning it is reasonable to expect the worst case scenario that future interventions are not effective.

We may take the approach that once *P_∞_* is attained this means the number of new reported outbreaks reaches a plateau too so the fraction lim*_t→∞_ I*_new_ = *Q_∞_* = *γrP_∞_*(1 − *P_∞_*) = *γP_∞_* per day. We may observe in data used and have *Q_∞_N* from master CQC list of care home^8^ whilst the recovery rate may be converted to the time to recovery *T* = 1/*γ*. Thus we may estimate *P_∞_* = *TQ_∞_*.

